# Monitoring the Risk of Type-2 Circulating Vaccine-Derived Poliovirus Emergence during Roll-Out of Type-2 Novel Oral Polio Vaccine

**DOI:** 10.1101/2024.10.16.24315616

**Authors:** Corey M. Peak, Hil Lyons, Arend Voorman, Elizabeth J Gray, Laura V Cooper, Isobel M Blake, Kaija Hawes, Ananda S Bandyopadhyay

## Abstract

**Background/Objectives:** Although wild poliovirus type 2 has been eradicated, prolonged transmission of the live-attenuated virus contained in the type-2 oral polio vaccine (OPV2) in under-immunized populations has led to emergence of circulating vaccine derived poliovirus type-2 (cVDPV2). The novel OPV2 (nOPV2) was designed to be more genetically stable and reduce the chance of cVDPV2 emergence while retaining comparable immunogenicity to the Sabin monovalent OPV2 (mOPV2). This study aims to estimate the relative reduction in the emergence risk due to use of nOPV2 instead of mOPV2.

**Methods:** Data on OPV2 vaccination campaigns from May 2016 to 1 August 2024 were analyzed to estimate type-2 OPV-induced immunity in children under 5 years of age. Poliovirus surveillance data were used to estimate seeding date and classify cVDPV2 emergences as Sabin- or novel-derived. The expected number of emergences if mOPV2 was used instead of nOPV2 was estimated accounting for the timing and volume of nOPV2 doses, the known emergence risk factors for emergence from mOPV2, and censoring due to the incomplete observation period for more recent nOPV2 doses.

**Results:** As of 1 August 2024, over 98% of the approximately 1.19 billion nOPV2 doses administered globally were in Africa. We estimate approximately 76 (95% confidence interval 69-85) index isolates of cVDPV2 emergences would be expected to be detected by 1 August 2024 if mOPV2 had been used instead of nOPV2 in Africa. The 18 observed nOPV2-derived emergences represent a 76% (74%-79%) lower risk of emergence by nOPV2 than mOPV2 in Africa. The crude global analysis produced similar results. Key limitations include the incomplete understanding of the drivers of heterogeneity in emergence risk across geographies and variance in the per-dose risk of emergence may be incompletely captured using known risk factors.

**Conclusions:** These results are consistent with the accumulating clinical and field evidence showing enhanced genetic stability of nOPV2 relative to mOPV2, and this approach has been implemented in near-real time to contextualize new findings during roll-out of this new vaccine. While nOPV2 has resulted in new emergences of cVDPV2, the number of cVDPV2 emergences is estimated to be approximately four-fold lower than if mOPV2 had been used instead.

## Introduction

The oral polio vaccine (OPV), developed by Albert Sabin in 1961, is an essential tool for the Global Polio Eradication Initiative (GPEI) and has been remarkably successful at providing both individual and population-level protection against poliovirus infection and poliomyelitis. The vaccine contains a live-attenuated virus which can produce a strong mucosal and humoral immune response following replication in the intestines and, once shed in stool, can yield secondary protection to contacts of the vaccine recipient. However, prolonged transmission of the vaccine virus in under-immunized populations can lead to the loss of attenuation and emergence of circulating vaccine-derived poliovirus (cVDPV).

Although wild poliovirus type 2 has been eradicated, cVDPV type 2 (cVDPV2) continues to spread and presents a Catch-22 challenge to poliovirus eradication, as the Sabin monovalent OPV type 2 (mOPV2) and trivalent OPV (tOPV) used to stop cVDPV2 outbreaks has also seeded emergent cVDPV2 outbreaks^1^. This challenge has grown dramatically following the April 2016 “Switch” from trivalent to bivalent OPV, which removed the type 2 component from routine OPV immunization^2^. A 2023 analysis of post-Switch cVDPV2 outbreaks in the African continent quantified the association between factors such as lower immunity and larger campaign size with an increasing risk of cVDPV2 emergence^3^.

The novel OPV2 (nOPV2) was designed in part to be more genetically stable and reduce the chance of cVDPV2 emergence while retaining comparable immunogenicity to mOPV2^4^. Among other design features, nOPV2 includes a stabilized domain V region. In mOPV2, the domain V is a key attenuation site which rapidly reverts to wild-type within an estimated mean time of 6.5 days^5^. Pre-clinical and clinical data on nOPV2 has shown substantially lower rates of reversion and neurovirulence compared to mOPV2^6^.

Based on pre-clinical and clinical trial data and the designation of polio as a Public Health Emergence of International Concern, nOPV2 received the first-ever Emergency Use Listing (EUL) by the World Health Organization (WHO) and its use rapidly scaled-up to millions of recipients through outbreak response under close field monitoring and enhanced surveillance to ensure safety, effectiveness, and genetic stability of nOPV2^7^. Viruses observed in the field can be distinguished as originating from nOPV2 or Sabin OPV2 based on evidence from whole genome sequencing including presence of any of the nOPV2 design features^8^.

As of 1 August 2024, 18 cVDPV2 emergences linked to nOPV2 had been reported, all from the African contient^9^. This study aims to estimate the relative reduction in emergence risk from the use of nOPV2 instead of mOPV2. In this paper, we compare cVDPV2 emergence events observed following nOPV2 use to the number of emergence events which could have been expected if mOPV2 was used instead.

## Materials and Methods

### Data

#### Vaccination campaigns

Since the ‘Switch’ from trivalent OPV (tOPV; types 1, 2, and 3) to bivalent OPV (bOPV; types 1 and 3) in April 2016, type-2 OPV has only been used in supplemental immunization activities (SIAs). Data from each SIA marked as completed and with a start date between May 2016 through July 2024 were downloaded on 1 August 2024 from the Polio Information System (POLIS) database maintained by the WHO. The number of OPV doses was estimated based on target population by campaign name as well as by province, and manually cross-checked against offline vaccination tracking data for recent campaigns. Total province population size was estimated assuming the under-5 SIA target population size represents 17% of the population demographic.

#### Immunity estimation

Type-2 OPV-induced immunity for children aged under 5 years was estimated in each district and month using a dynamic model assuming 80% per-campaign coverage^10^. To estimate the pre-campaign immunity in each province, we calculated the population-weighted average district immunity for the month prior to each campaign start date.

#### Virus surveillance

Poliovirus surveillance depends on isolation of the virus from human stool specimens (from acute flaccid paralysis surveillance or contact sampling) or sewage samples from environmental surveillance (ES). Standard guidelines include up to 10 days in cell culture followed by Sanger sequencing. Data on polioviruses were downloaded from POLIS and cVDPV2 Emergence groups were assigned as Sabin-derived or nOPV2-derived based on classification by whole genome sequencing^8^. Provinces were classified as having ES if there was at least one ES result in the past year.

### Analysis

#### Estimating seeding date

The mean seeding date for cVDPV2 was estimated based on the number of nucleotide changes in the VP1 region of the index isolate compared with mOPV2, using a mutation clock estimated as two instantaneous mutations then 9 changes per year following a Poisson process^5^. cVDPV2 emergence groups with index viruses with mean estimated seeding date after April 2016 were counted as emergences seeded after the Switch.

#### Estimating time to detection

To account for the time lag between the use of OPV and the detection of any subsequent emergences, we estimated the distribution of emergence waiting times and reporting waiting times (surveillance lags). The emergence waiting time distribution is defined as the time from the potential seeding event (ie, the OPV2 SIA) to the index virus date, and was estimated using a mixture model of mutations given OPV2 exposures in SIAs and surveillance type. The viral age of the index isolate in each emergence group was subtracted from the virus date (ie, AFP onset date or ES collection date) to generate a probabilistic time period during which seeding may have occurred. OPV2 exposure was estimated using the SIA starting date, number of OPV2 doses, and distance to district of emergence based on a radiation model of decay over space^11^. Emergence waiting time was estimated with a log-normal distribution for all emergences, and again ignoring ES to reflect waiting times in a setting with only AFP surveillance.

The reporting waiting time is defined as the time from virus date (ie, AFP onset or ES collection date) until notification of VP1 viral sequence. Focusing on cVDPV2 detections after 1 May 2016, we estimated the waiting time using a mixture of gamma distributions with random effects per country for the mean and shape parameters. We ignored isolates that took longer than 1 year to report, which are likely de-prioritized samples as compared with samples of potential epidemiological importance such as potential new emergences. Since the sequencing lag begins after the emergence waiting time, the convolution of the distributions was calculated to construct the time to detection of the index isolate for a new emergence group.

#### Estimating number of emergences expected

First, we performed a global crude analysis based on observed emergence rate per post-switch Sabin OPV2 dose. Scaling this rate by the number of nOPV2 doses used provides a total number of emergences that would be expected if each nOPV2 SIA during the period from March 2021 through July 2024 had been replaced with mOPV2. 95% confidence intervals were constructed based on the Poisson distribution. The time to detection distribution following each nOPV2 SIA was then scaled such that the area under the curve equaled the expected number of emergences from that SIA. The expected number of emergences detected by a given day can then be estimated as the cumulative sum up to that day.

Given the vast majority of nOPV2 doses have been administered in the African continent, and previous analyses have estimated the relationship between campaign size, pre-existing immunity, and the consequent cVDPV2 emergence rate from Sabin OPV2, we performed an adjusted analysis focused on AFR. Using the framework and fitted parameters *θ*_*size*_ and *θ*_*u*5_ reported by Gray et al^3^, we estimated 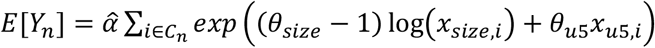, where *E*[*Y*_*n*_] is the expected number of emergences from a set *C*_*n*_ of EUL-period nOPV2 SIAs, each with size *x*_*size,i*_ and pre-campaign immunity *x*_*u*5,*i*_. The scaling factor 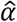 is estimated based on the *Y*_*s*_ observed emergences from a set *C*_*s*_ of post-Switch mOPV2 or tOPV SIAs according to 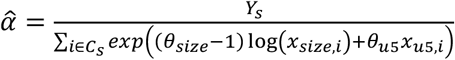. To account for parameter uncertainty, *E*[*Y*_*n*_] was calculated for each of 500 samples from the joint posterior (*θ*_*size*_, *θ*_*u*5_) Markov Chain Monte Carlo output. The 95% prediction interval was constructed as the 2.5^th^ and 97.5^th^ percentiles. As per the crude analysis, the number of expected emergences was distributed based on the time to detection.

## Results

### Description of post-Switch OPV2 use

Between 1 May 2016 and 1 August 2024, 127 million doses of tOPV, 551 million doses of mOPV2, and 1.19 billion doses of nOPV2 have been administered (Table 1). Use of nOPV2 has increased rapidly since its first use in March 2021 and represented 100% of total OPV2 use since Q2 2023 (**Figure 1**). While global crude results are presented, given that 1.17 of the 1.19 billion nOPV2 doses (98%) used has been in Africa, the main results in this paper focus on emergence risk in Africa, with spotlights on Nigeria, Democratic Republic of the Congo (DRC), and other priority countries.

**Table 1.** Use of OPV2 and subsequent emergences first detected in Africa or elsewhere (1 May 2016 to 1 August 2024)

|  | Country<sup>i</sup> | Vaccine doses (Million)<sup>ii</sup> |  | cVDPV2 Emergences<sup>iii</sup> |  |
| --- | --- | --- | --- | --- | --- |
|  |  | Sabin OPV2 | nOPV2 | Derived from Sabin OPV2 | Derived from nOPV2 |
| **Africa** |  | **483** | **1,170** | **59** | **18** |
|  | *Nigeria* | *163* | *657* | *10* | *2* |
|  | *DRC* | *53* | *89* | *17* | *5* |
| **Elsewhere** |  | **196** | **20** | **21** | **0** |
| **Total** |  | **679** | **1,190** | **80** | **18** |

<sup>i</sup> Select countries of interest shown.
<sup>ii</sup> Data: POLIS campaigns marked completed with start dates between 1 May 2016 and 1 August 2024.
<sup>iii</sup> Emergences first detected in location with mean estimated seeding date after 1 May 2016.

**Figure 1.**
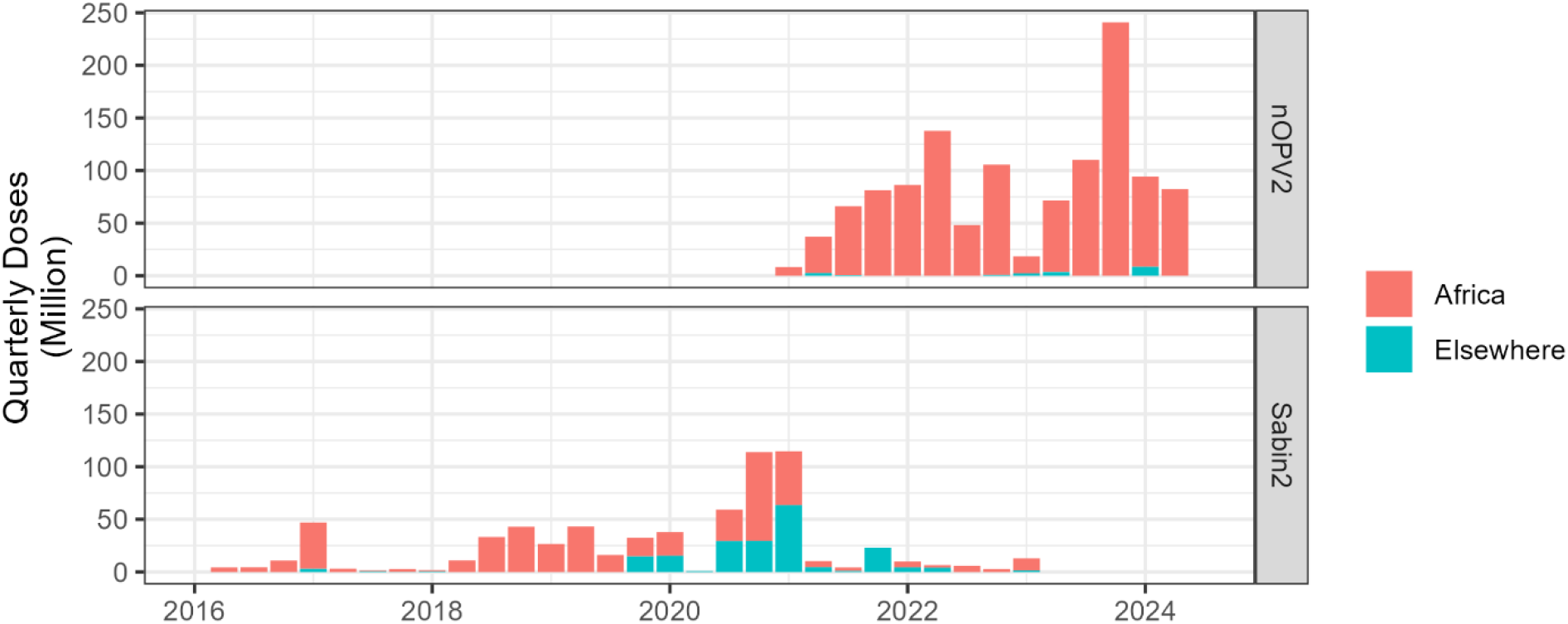
Quarterly number of OPV2 doses by vaccine product.

The median target population size is smaller for post-Switch campaigns in Africa with Sabin OPV2 (646,204 [IQR 266,388 – 1,690,858]) than nOPV2 (2,766,913 [IQR 1,195,384 – 6,159,151]) (p<0.001)(**Figure 2**). Estimated pre-campaign type-2 mucosal immunity among children aged under 5 years is similar for post-Switch campaigns in Africa with Sabin OPV2 (median 0.83 [IQR 0.75 – 0.91]) and nOPV2 (median 0.86 [IQR 0.76 – 0.93]) (p=0.15) (**Figure 2**), though nOPV2 was generally used in lower immunity contexts than mOPV2 in DRC.

**Figure 2.**
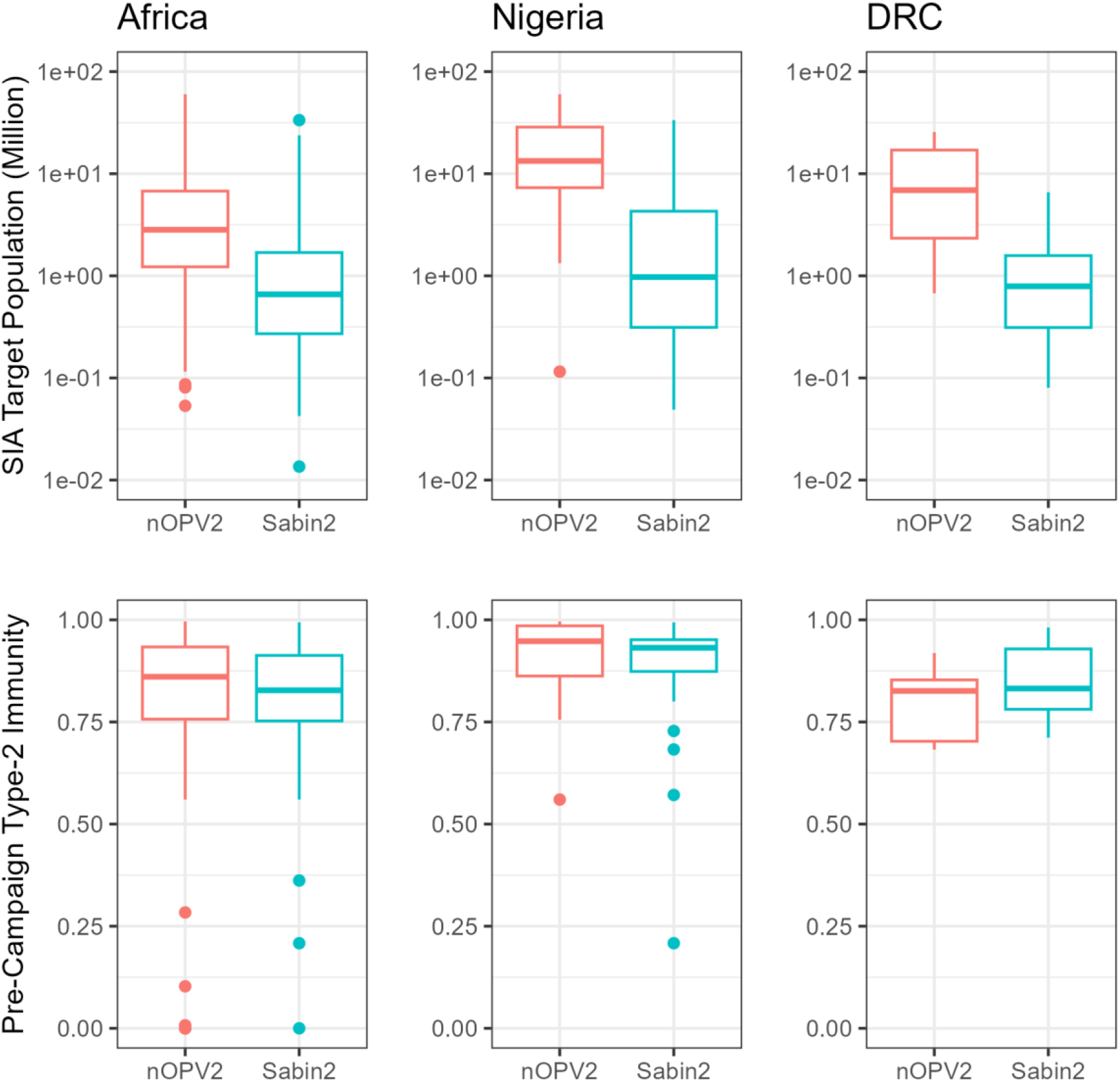
Boxplots of SIA Size (top, note log axis) and pre-campaign immunity (bottom) for the Africa, Nigeria, and DRC.

### Observation of post-Switch cVDPV2 emergences

87 cVDPV2 emergences derived from Sabin OPV2 viruses have been reported since 1 May 2016, of which 80 have an estimated seeding date after the “Switch” in April 2016. 59 of these post-Switch Sabin OPV2 emergences were first detected in Africa(**Table 1**). The crude observed seeding risk per 100 million Sabin OPV2 doses is 12.2 in Africa and 10.7 elsewhere.

18 cVDPV2 emergences derived from nOPV2 have been reported, all from Africa(**Figure 3**). The crude observed seeding risk per 100 million nOPV2 doses is 1.5 in Africa; however, the process for emergence and detection by surveillance creates a time lag such that emergences due to more recent campaigns (largely nOPV2) are only partially observed.

**Figure 3.**
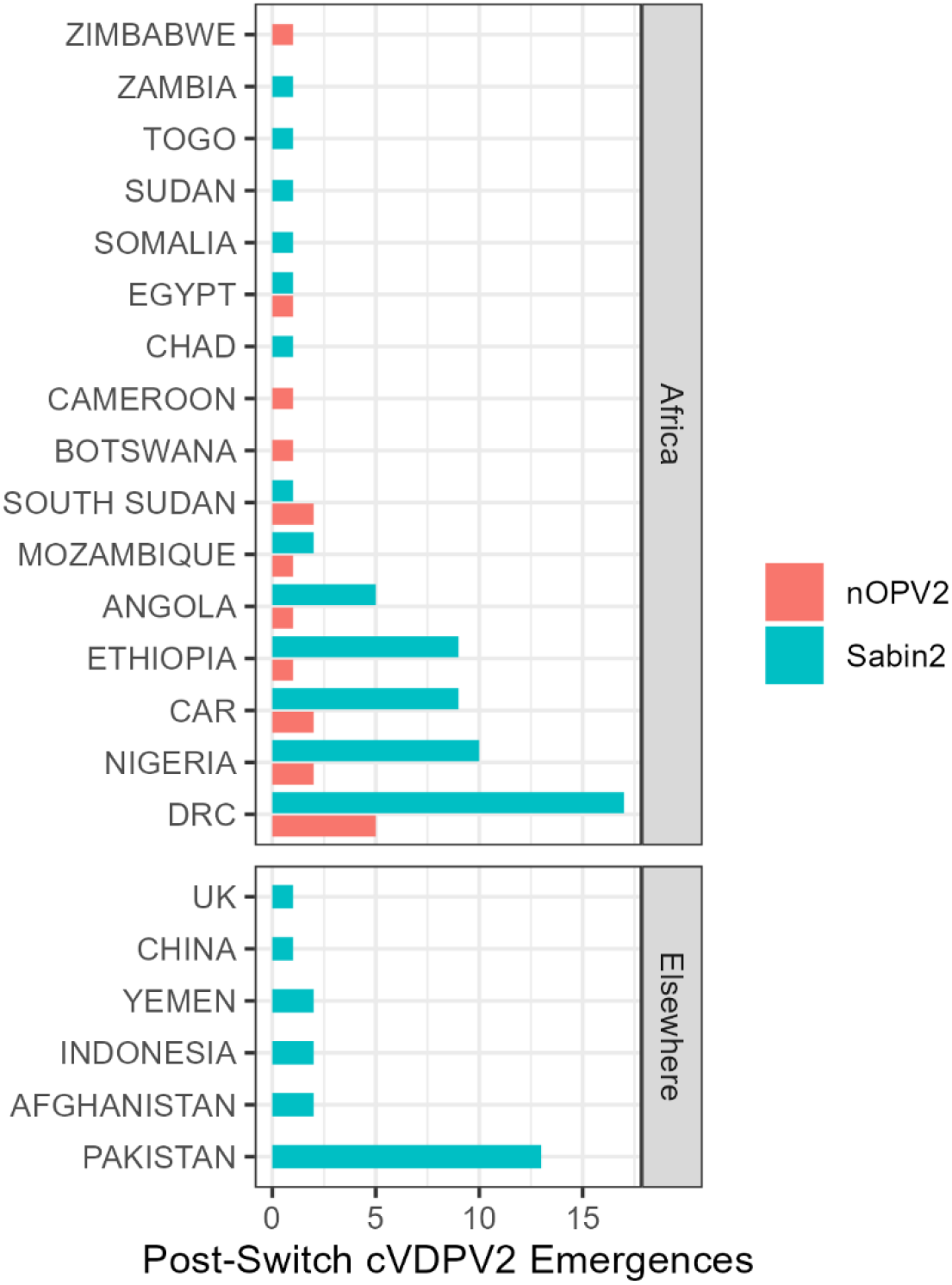
Countries detecting index isolate of cVDPV2 emergences seeded after April 2016.

### Estimating emergence expectation for nOPV2

Accounting for estimated viral age as well as the timing and proximity of SIAs, the onset date for the index virus of an estimated 60% of cVDPV2 emergences was detected by AFP surveillance within 12 months of the likely SIA seeding event; where there is AFP and environmental surveillance, 94% of index viruses were collected within 12 months of the SIA (**Appendix Figure 1**). Time from virus date until reporting of the viral sequence was estimated for each country; for example, 50% of sequences were reported within 56 days of virus date in Nigeria and 77 days in Democratic Republic of the Congo (**Appendix Figure 2**).

### Crude Global Analysis

A crude global emergence rate of 11.8 cVDPV2 emergence per 100 million Sabin OPV2 doses was estimated based on 80 cVDPV2 emergences derived from 679 million post-switch Sabin OPV2 doses. Applying this crude rate to the 1.19 billion nOPV2 doses used, an estimated 140 (95% confidence interval 117-164) cVDPV2 emergences would be expected if Sabin OPV2 had been used instead of nOPV2. However, not all of these emergences would be observable by August 2024. Applying the estimated emergence waiting time and reporting waiting time distributions, the index isolate for 93 (75-113) of these emergences would be expected to have been discovered by 1 August 2024. The 18 cVDPV2 emergences from nOPV2 that have been discovered by then represent an estimated 81% (76%-84%) lower risk of emergence by nOPV2 than Sabin OPV2.

### Adjusted Africa Analysis

An adjusted analysis was performed for the African continent, where the vast majority of nOPV2 doses have been used and where additional analyses on emergence risk factors has been performed^3^. An estimated 123 (115–135) cVDPV2 emergences would have been expected if Sabin OPV2 had been used instead of nOPV2 given total nOPV2 use in AFR during the period from March 2021 to 1 August 2024. Applying the estimated waiting time and reporting time distributions, the index isolate for 76 (69-85) of these emergences would be expected to have been discovered by 1 August 2024 (**Figure 4**). The 18 cVDPV2 emergences from nOPV2 that have been observed by then represent an estimated 76% (74%-79%) lower risk of emergence by nOPV2 than mOPV2 in Africa.

**Figure 4.**
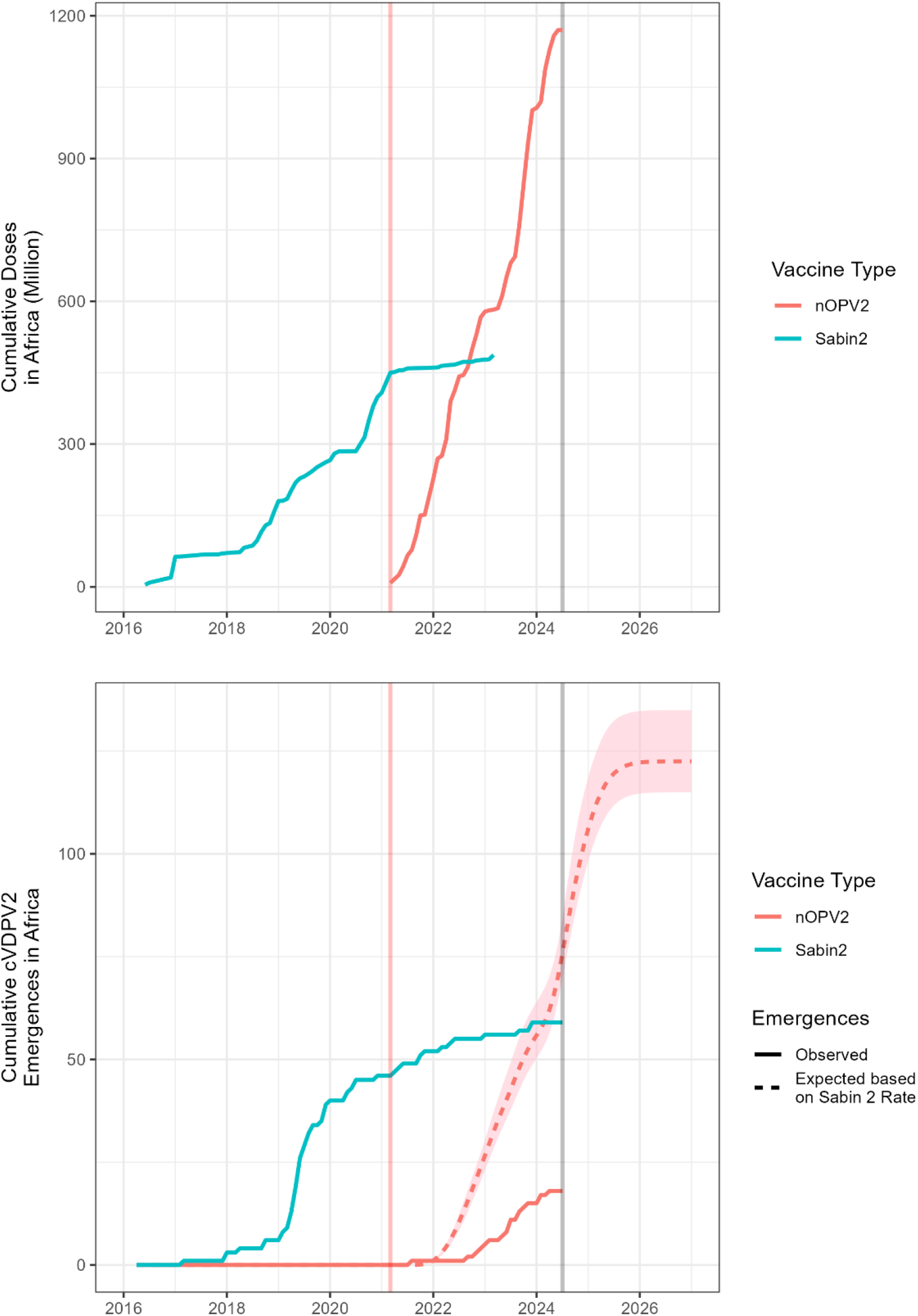
(a) Cumulative doses of Sabin OPV2 and nOPV2 in Africa; (b) Cumulative observed and expected cVDPV2 emergences derived from Sabin OPV2 and nOPV2 in Africa, by virus date of index isolate. Vertical lines indicate date of first nOPV2 use (red) and August 2024 (grey).

### Sensitivity analyses

There remains residual uncertainty in emergence risk by location after considering known risk factors^3^. The main results narrow the scope to Africa, representing 98% of nOPV2 use, though sub-regional geographic heterogeneity in emergence risk remains. In Nigeria, 10 cVDPV2 emergences derived from 163 million doses of Sabin OPV2 and 2 cVDPV2 emergences from 657 million doses of nOPV2 have been observed (**Table 1**). Estimating emergence expectations for Nigeria separately, approximately 24 (23-25) cVDPV2 emergences could be expected by 1 August 2024 if mOPV2 were used instead of nOPV2, representing an estimated 92% (91%-92%) lower risk of emergence by nOPV2 than mOPV2 (**Appendix figure 3**). Conversely, estimating emergence expectations for DRC separately, the observed 5 cVDPV2 emergences derived from nOPV2 represent a 57% (48%-63%) decrease compared with the 12 (10-13) cVDPV2 emergences that could be expected by 1 August 2024 if mOPV2 were used instead (**Appendix figure 4**).

While 59 cVDPV2 emergences derived from Sabin OPV2 have been observed since 1 May 2016 in Africa, a cluster of 4 emergences from April-June 2019 in two provinces in Angola have been hypothesized to have related origins, and likewise for a cluster of 5 emergences in May 2019 from two provinces in CAR^12^. By treating the Angolan cluster as a single emergence event, and likewise for the CAR cluster, we can repeat the analysis based on a lower emergence risk for mOPV2 and estimate that 67 (61-75) cVDPV2 emergences could be expected by 1 August 2024 if mOPV2 had been used instead of nOPV2 in Africa, representing a 73% (71%-76%) lower risk of emergence by nOPV2 than mOPV2. However, consolidating these clusters is expected to influence the parameters of the emergence risk analysis in a way not accounted for here.

## Discussion

These results support the growing body of evidence from clinical and field studies demonstrating nOPV2 is more genetically stable and less likely to revert and lead to cVDPV2 emergence than Sabin mOPV2. The risk of cVDPV2 emergence is estimated to be five-fold lower following nOPV2 use than Sabin mOPV2. Importantly, the risk of cVDPV2 emergence can be further reduced by implementing high-quality campaigns to prevent the circulation, and hence reversion, of all types of polioviruses. These results support the use of nOPV2 for response to cVDPV2 outbreaks and as a critical tool on the path to polio eradication. Previous work provided preliminary estimates of a ten-fold reduction in emergence risk for nOPV2 compared with Sabin OPV2 based on a smaller, earlier set of data and before accounting for times to cVDPV2 emergence and reporting which are included here to address the partially-observed nature of real-time monitoring of emergence risk^13^.

Encouraging field performance of nOPV2 underscores importance of the tool for containing spread of current outbreaks while carrying a lower risk of seeding new emergences. Risk assessments for scoping outbreak response campaigns must still be mindful of emergence risk, though the lower emergence risk with nOPV2 than Sabin OPV2 shifts the balance towards larger campaigns to stop spread. However, the seeding risk is not zero, underscoring the importance of known ways to reduce the emergence risk, including implementing high-coverage SIAs in rapid succession^14^.

This work is subject to limitations including the following. First, the risk of cVDPV2 emergence is heterogeneous and its drivers are incompletely understood and modeled. For example, further study is warranted to characterize the role of non-polio enteroviruses (NPEV) as recombination partners on the critical pathway to cVDPV2 emergences and spatiotemporal prevalence of these NPEV species^6^.

Immunity estimation may only partially address variation in emergence risk across repeated SIAs in the same location; due to the large volume of nOPV2 used in Nigeria beyond the second round, emergence risk may attenuate more, or less, quickly than immunity estimates capture alone. Further, the model does not capture differences in emergence risk by relative campaign frequency. E.g. all four nOPV2-derived emergences in DRC appear consistent with seeding during the first nOPV2 round in a province which had no OPV2 use within 2-4 years, and which was followed by a second round approximately 12 weeks later, thereby allowing time for nOPV2 viruses to circulate and revert. Reviewing such evidence, the WHO SAGE in 2023 reiterated the recommendation to plan for a second SIA no later than 4 weeks after the first SIA in a location^14^.

Second, the emergence waiting time distribution is fitted to data on cVDPV2 emergences derived from Sabin OPV2, and comparisons to this waiting time assume that the waiting time for nOPV2 would be similar. We posit this is a conservative assumption, since the biological pathway to cVDPV2 emergence via recombination (the predominant reversion pathway for nOPV2) is expected to occur more quickly than the pathway via accumulation of point mutations (plausibly the predominant reversion pathway for Sabin OPV2^5^). Furthermore, changes in surveillance sensitivity with time, especially increase in ES^15^, may result in faster surveillance more recently. Therefore, if any cVDPV2 emergences occur at all, one may expect to see them more quickly following campaigns with nOPV2 than Sabin OPV2, and therefore the Sabin OPV2 comparator in this analysis may provide a conservative safety margin in this respect.

Furthermore, definition of a cVDPV2 emergence group requires an index isolate and linked confirmatory isolate. However, the median time from index-to-confirmatory isolate for post-switch cVDPV2 emergence groups in Africa is 27 days, suggesting this introduces a lag of approximately one month.

Third, the Sabin OPV2 comparator assumes that an equal number of Sabin OPV2 doses would have been used instead of nOPV2, and therefore that no additional doses of nOPV2 were needed for a given response compared with Sabin OPV2. The growing evidence base supports similar immunogenicity, efficacy, and field effectiveness of nOPV2 and mOPV2^9,16–19^.

Fourth, non-differential measurement error can introduce noise for several data fields. The number of doses used in each campaign are estimated based on target population for the SIA, the quality of which varies by location. Due to limited surveillance sensitivity, including the low case-to-infection ratio for polio, the location where an emergence is first detected may not be the location where the virus was first circulating and reverted. This introduces noise into understanding the risk factors for where emergence occurs, and not just where emergence is first detected (which may be a function of higher relative surveillance sensitivity or transmission proclivity). Districts with an ES site were assumed to follow the ES+AFP time to discovery distributions, which does not account for potential differences in site sensitivity^20^. Estimation of immunity depends on an assumed 80% per-round vaccination coverage with random missingness, while in reality, sub-district pockets of lower coverage are expected to be important drivers of transmission and potentially emergence. In the case studies of Nigeria and DRC, quality of each campaign is uncertain, with potentially meaningful implications on the differences observed in these countries.

Further work to understand the risk factors for emergence would help to inform this analysis and support generalization of novel OPV types 1 and 3 under development. Furthermore, while lower emergence risk is unequivocally better, a key question remains - how low emergence risk must be to achieve eradication under various programmatic implementation conditions.

## Conclusions

While nOPV2 has resulted in new emergences of cVDPV2, the number of cVDPV2 emergences is estimated to be approximately four-fold lower than if mOPV2 had been used instead. These results are consistent with the accumulating clinical and field evidence showing enhanced genetic stability of nOPV2 relative to mOPV2. By accounting for detection timelines, this approach has been and can continue to be updated in near-real time to contextualize new findings during the roll-out of nOPV2.

## Data Availability

Code available at: https://github.com/peakcm/polio

## Appendix

**Appendix figure 1.**
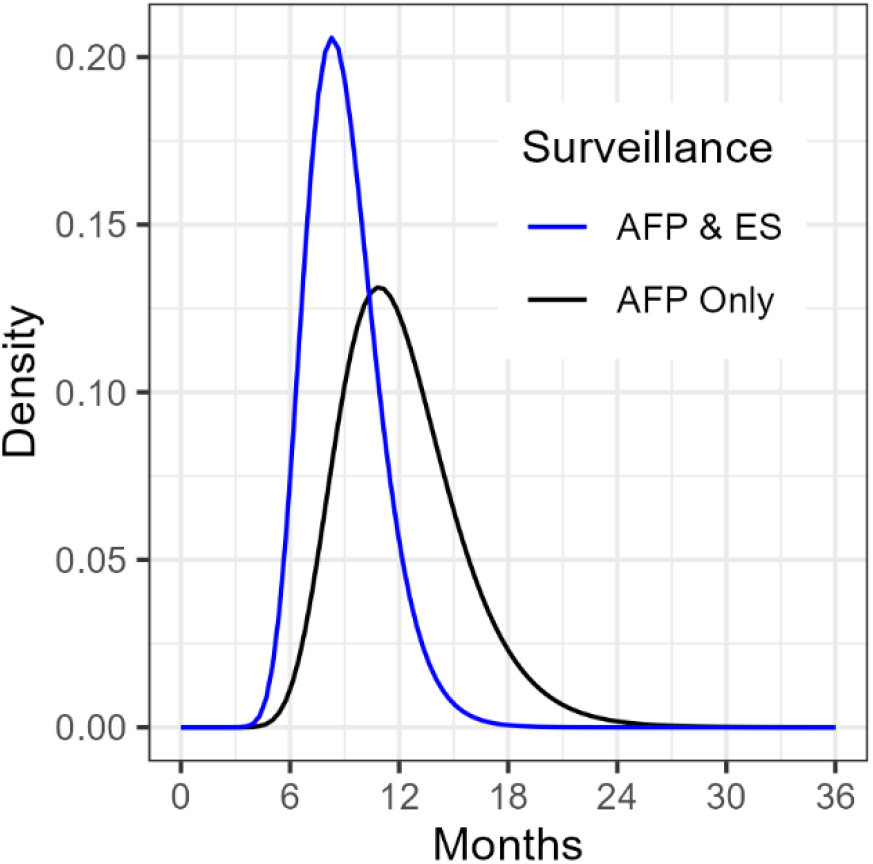
Emergence Waiting Time distributions.

**Appendix figure 2.**
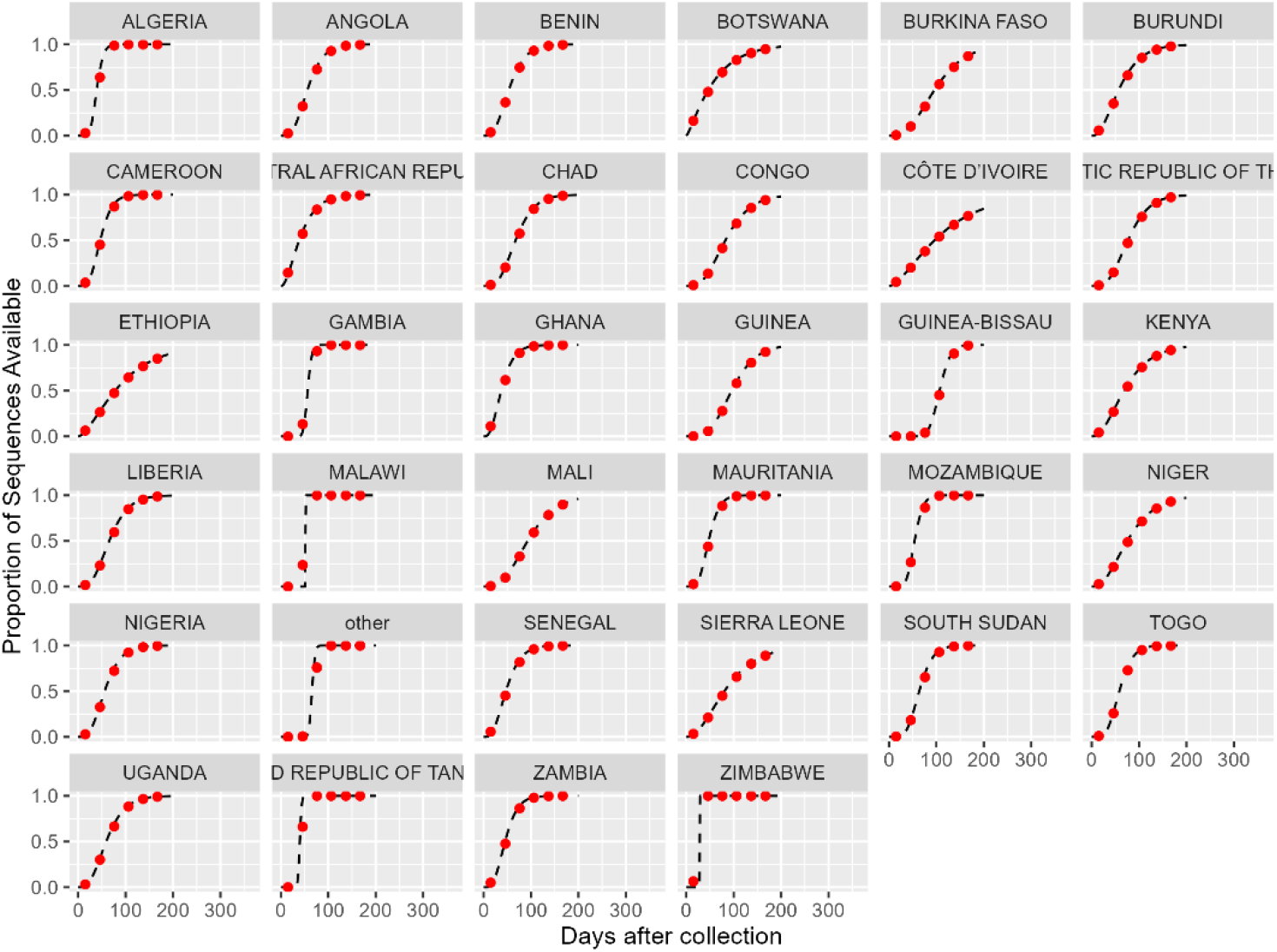
Reporting Waiting Time distributions. *“Other” is other countries globally. Red dots indicate monthly mean values, at monthly midpoints*.

**Appendix figure 3.**
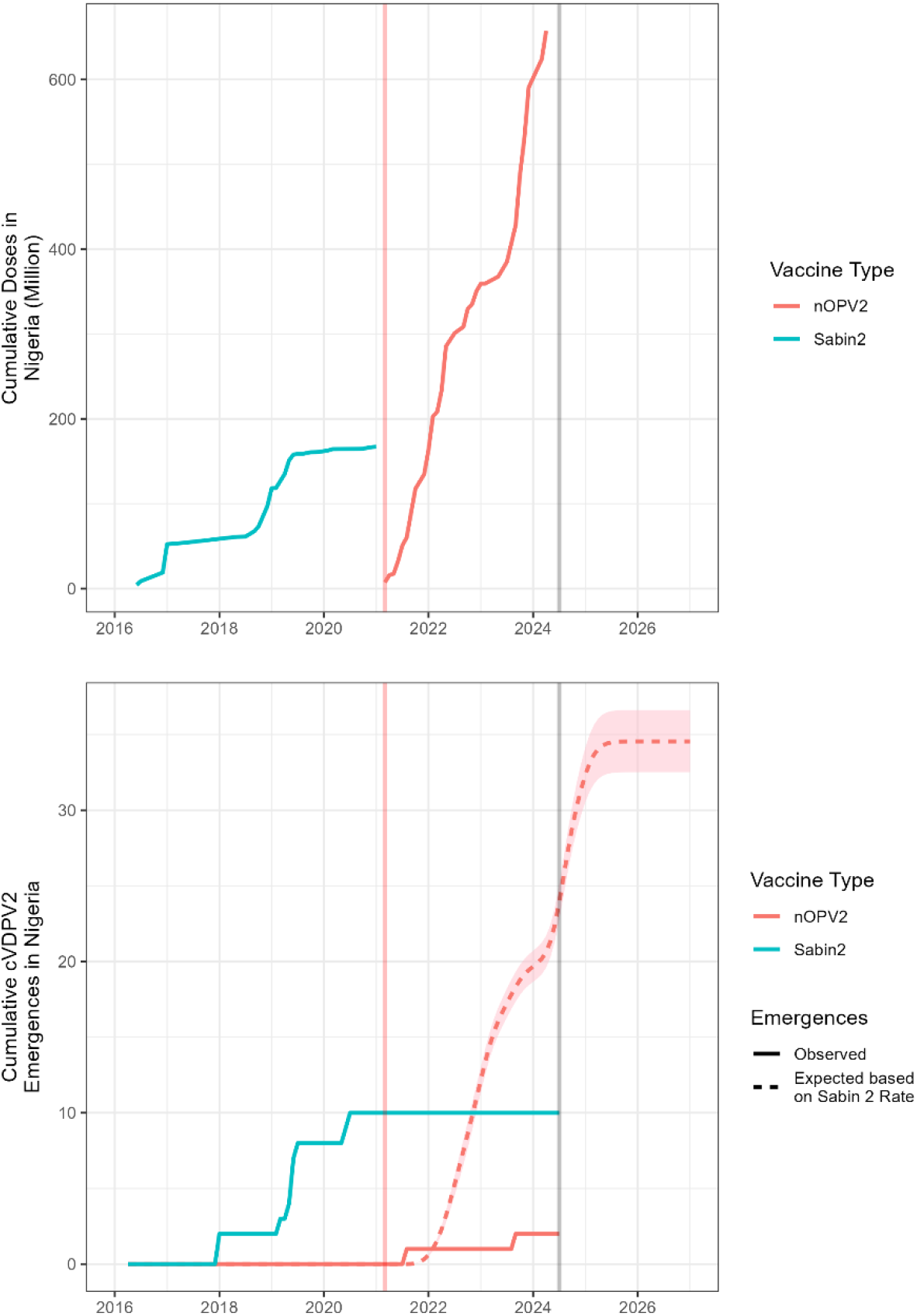
(a) Cumulative doses of Sabin OPV2 and nOPV2 in Nigeria; (b) Cumulative observed and expected cVDPV2 emergences derived from Sabin OPV2 and nOPV2 in Nigeria, by virus date of index isolate. Vertical lines indicate date of first nOPV2 use (red) and April 2024 (grey).

**Appendix figure 4.**
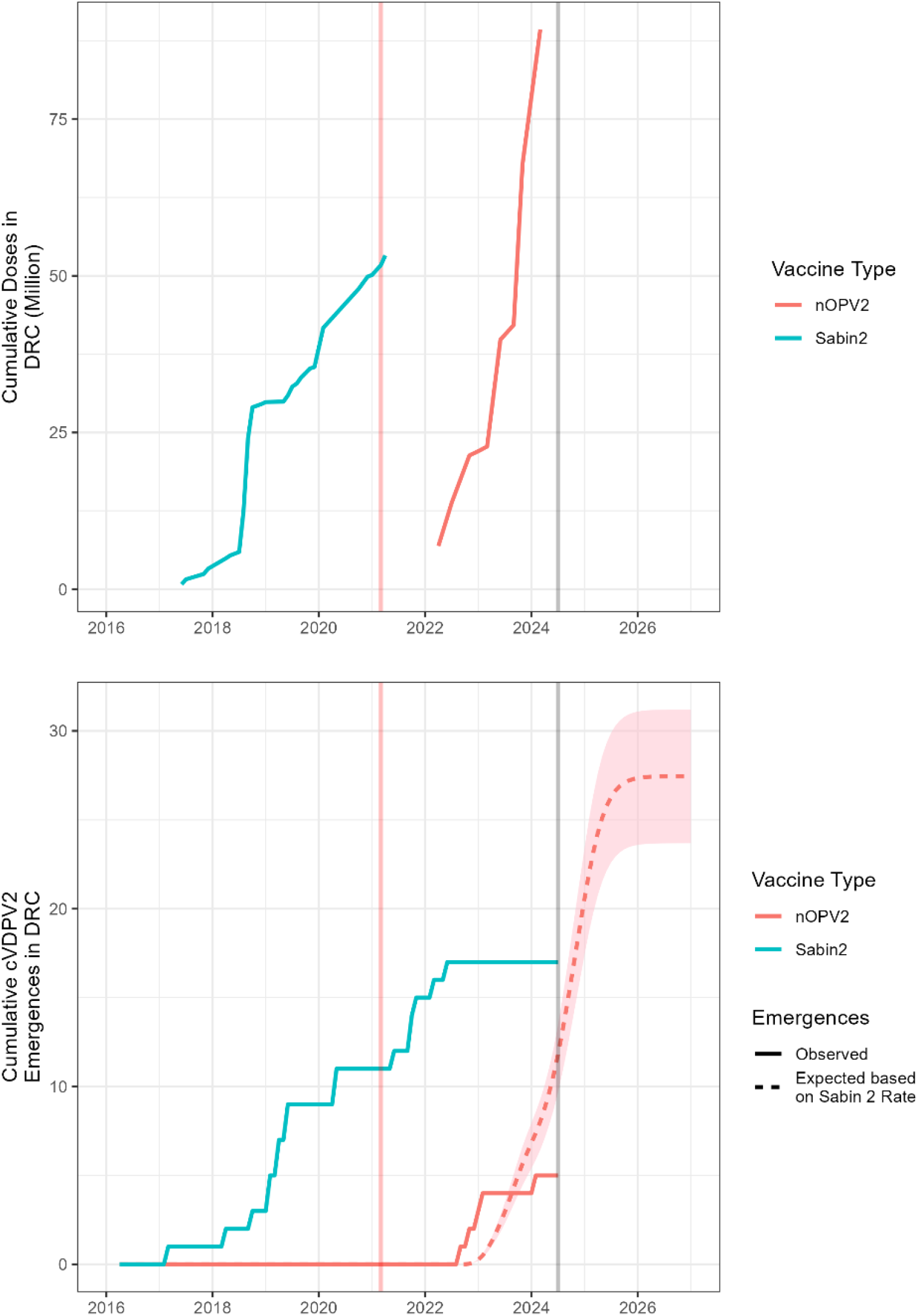
(a) Cumulative doses of Sabin OPV2 and nOPV2 in DRC; (b) Cumulative observed and expected cVDPV2 emergences derived from Sabin OPV2 and nOPV2 in DRC, by virus date of index isolate. Vertical lines indicate date of first nOPV2 use (red) and April 2024 (grey). Code available here: https://github.com/peakcm/polio

